# Second round statewide survey for estimation of the burden of active infection and anti-SARS-CoV-2 IgG antibodies in the general population of Karnataka, India

**DOI:** 10.1101/2021.08.10.21261842

**Authors:** M Rajagopal Padma, Prameela Dinesh, Rajesh Sundaresan, Siva Athreya, Shilpa Shiju, Parimala S Maroor, R Lalitha Hande, Jawaid Akhtar, Trilok Chandra, Deepa Ravi, Eunice Lobo, Yamuna Ana, Prafulla Shriyan, Anita Desai, Ambica Rangaiah, Ashok Munivenkatappa, S Krishna, Shantala Gowdara Basawarajappa, HG Sreedhara, KC Siddesh, B Amrutha Kumari, Nawaz Umar, BA Mythri, KM Mythri, Mysore Kalappa Sudarshan, Ravi Vasanthapuram, Giridhara R Babu

## Abstract

**Objective:** The second round of the serial cross-sectional sentinel-based population survey to assess active infection, seroprevalence, and their evolution in the general population across Karnataka was conducted. Additionally, a longitudinal study among participants identified as COVID-19 positive in the first survey round was conducted to assess the clinical sensitivity of the testing kit used.

**Methods:** The cross-sectional study of 41,228 participants across 290 healthcare facilities in all 30 districts of Karnataka was done among three groups of participants (low, moderate, and high-risk). Consenting participants were subjected to real-time reverse transcription-polymerase chain reaction (RT-PCR) testing, and antibody (IgG) testing.

**Results:** Overall weighted adjusted seroprevalence of IgG was 15.6% (95% CI: 14.9–16.3), crude IgG prevalence was 15.0% and crude active prevalence was 0.5%. Statewide infection fatality rate (IFR) was estimated as 0.11%, and COVID-19 burden estimated between 26.1 to 37.7% (at 90% confidence). Clinical sensitivity of the IgG ELISA test kit was estimated as ≥38.9%.

**Conclusion:** The sentinel-based population survey helped identify districts that needed better testing, reporting, and clinical management. The state was far from attaining natural immunity during the survey and hence must step up vaccination coverage and enforce public health measures to prevent the spread of COVD-19.

## INTRODUCTION

The COVID-19 pandemic has spread globally and affected 2.58% of the population, with a case fatality rate of 2.12% as of 4 August 2021. In India alone, 31.8 million people were diagnosed with COVID-19 with a case fatality rate of 1.44%.[1] As the pandemic continues to progress, most countries from South Asia to Europe have seen a more severe second wave.[2] While data on reported cases, deaths, and testing drive the short-term management of the pandemic, given the high rate of asymptomatic infection in the population[20] that may go undetected, it is important to estimate active infection and seroprevalence in the general population for better matching of public health responses to the actual state of the pandemic.

Evidence from nationwide surveys in India, conducted by the Indian Council of Medical Research (ICMR), reported that the antibodies to SARS-CoV-2 were detected in 0.73% population during May-June 2020 (first round)[13], in 6.6% during August-September 2020 (second round)[14] as daily cases and deaths peaked in the country, and in 24.1% of adults surveyed and 27.2% of 10 to 17-year-olds surveyed during December 2020 - January 2021 (third round).[22] Maharashtra, Kerala, Karnataka, and Tamil Nadu reported the highest number of confirmed cases at the state level.[16] Seroprevalence varied from 0.13% in Kerala [7] in an early study ending 31 May 2020 to 31.6% in Tamil Nadu[12] in a study ending 30 November 2020. In the first round of the survey in Karnataka, the estimated total burden was 27.7% as of 16 September 2020[17], while a higher prevalence of 39.6% was reported in select households.[15] All states in India, including Karnataka, showed a decreasing trend from mid-October 2020 to January 2021. Further, studies have found declining IgG levels in the general population.[18],[19],[26],[27] Therefore, it is important to assess the active infection and seroprevalence in the population periodically.

Serial cross-sectional sentinel-based population surveys [17], conducted at different time points, provide insights on the epidemiological trend of infection spread. The cross-sectional nature provides a snapshot of the state of the pandemic across the survey region. The sentinel nature enables rapid and easier implementation. The serial nature ensures high-quality data from the same locations and population segments for capturing trends.

We conducted such a survey across Karnataka for the second time. Given the significant variation in IgG titres in the infected population [26][27][28][29][30] and the evidence of declining levels of IgG in the general population[18][19][31], we also conducted a longitudinal study among participants who were identified as COVID-19 positive in the September 2020 first round of our survey to assess the *clinical sensitivity* of the testing kit.

## METHODS

### The survey

We followed a protocol similar to the first round (Round 1) in September 2020[17] to estimate the fraction of the population with active infection and IgG antibodies at the time of the survey.

#### Setting

The study was conducted in all 30 districts of Karnataka and eight administrative zones of the Bengaluru metropolitan area. This subdivision led to a total of 38 units across the state. Health facilities were selected based on geographical representation, feasibility, ease of recruitment and were the same as in Round 1[17].

#### Sampling frame

The study sampled three population groups: low-, moderate-, and high-risk groups. The low-risk group comprised pregnant women presenting for a regular check-up at the ante-natal care (ANC) clinic and attenders of patients coming to the outpatient department in the healthcare facilities. The moderate-risk group comprised people with high contact in the community, e.g., bus-conductors, vendors at the vegetable markets, healthcare workers, *pourakarmikas* (waste-collectors), and individuals in congregate settings (such as markets, malls, retail stores, bus stops, railway stations, and hotel staff). The high-risk group, or more appropriately the vulnerable group, comprised the elderly and persons with comorbid conditions.

#### Sample size

For a margin of error of 0.05 and a 95% confidence level, taking design effect to be 3, assuming 32.3% prevalence, which is 5% more than the total burden estimated in Round 1,[17] the minimum required sample size was 1050 per unit[24] or 39,900 across the 38 units. The 1050 samples per unit were divided equally (350 each) among the three risk groups and were further divided equally among the risk sub-groups.

#### Inclusion and exclusion criteria

We included all adults ≥18 years. We excluded those already diagnosed with SARS-CoV-2 infection, those unwilling to provide a sample for the test or consent, those who had received vaccination for COVID-19, and those who already participated in Round 1.

#### Data collection

We obtained written informed consent from all participants prior to recruitment. We then collected the meta-data of all consenting participants (demographic details, comorbidities, and symptoms suggestive of COVID-19 in the preceding one month).

#### Sample collection and lab tests

For the reverse transcription-polymerase chain reaction (RT-PCR) test, we collected nasopharyngeal/oropharyngeal swabs. We used the current ICMR protocol for sample collection, cold-chain transport, and laboratory analysis and tested them through the ICMR-approved testing network. For IgG antibody testing, we collected 4 ml of venous blood, centrifuged it, transported the serum to the laboratory while maintaining a cold chain, and detected SARS-CoV-2-specific IgG antibodies using a commercial, ICMR-approved, ELISA-based test kit (Covid Kavach Anti SARS-Cov-2 IgG antibody detection ELISA, Zydus-Cadila, India) [25] following the manufacturer’s instructions. Results were declared positive or negative based on the cut-off value of optical densities obtained with the positive and negative controls provided with the kit. Supplementary Figure 1 contains the schema for the laboratory tests conducted, while Supplementary Figure 2 shows the survey algorithm.

### Longitudinal study for antibody waning

A longitudinal study to assess the clinical sensitivity of the test kit, in view of antibody waning, was also conducted.

#### Setting, sampling frame, and sample size

In Round 1, around 4582 out of 15939 participants from all units tested positive on at least one of the tests (the rapid antigen test, which was conducted in Round 1 but not in Round 2, the RT-PCR test, and the antibody test). Of these, 4420 participants from all risk groups with unambiguous meta-data were selected for the longitudinal study expecting that 10-20% would agree to participate.

#### Exclusion criteria

We excluded those with a breakthrough infection (after Round 1), those that were vaccinated, and those that did not provide informed consent.

#### Data collection, sample collection, and lab tests

We obtained written informed consent from all participants prior to the study. As indicated above, we collected 4 ml of venous blood from each consenting participant, centrifuged it, transported the serum to the laboratory while maintaining a cold chain, and detected SARS-CoV-2-specific IgG antibodies using the same ELISA-based test kit (Zydus-Cadila)[25].

### Ethical Considerations

The Institutional Ethics Committee (IEC) of the Indian Institute of Public Health – Bengaluru campus reviewed and approved the study (vide. IIPHHB/TRCIEC/174/2020) and the subsequent change of protocol to perform the longitudinal study (vide PHFI/IIPH-BLR/076/2020-21). We informed the participants of the purpose of the surveys, how the samples would be taken and requested them to respond to the screening questions. After obtaining informed consent, we noted basic demographic details, exposure history, symptoms observed in the previous month, and clinical history. Participants’ test results were shared with them by the concerned healthcare facility.

### Statistical Analysis

IgG prevalence was defined as the fraction of the sampled population with *detectable* IgG antibodies; active infection fraction was defined as the fraction of the sampled population who test positive on the RT-PCR test, and total prevalence of COVID-19 was defined as the fraction of the sampled population with either detectable IgG or active infection.

For the estimation of IgG prevalence, active infection fraction, total prevalence, confidence intervals, and the odds ratios, we followed the method as outlined in [17]. For predicting IgG prevalence based on co-morbidities and other factors, we used logistic regression.

The longitudinal study was used to estimate the clinical sensitivity of the ELISA kit. Considering the significant lapse of time between Round 2 (end-date 18 February 2021) and the longitudinal study (end-date 11 May 2021), we only obtained a lower bound on the clinical sensitivity. This yields an upper bound on the total prevalence.

## RESULTS

### The second-round serial cross-sectional sentinel-based population survey

The statewide survey was carried out in 290 healthcare facilities spread across Karnataka from 25 January to 18 February 2021. Of the 44539 people approached, 115 refused, and 3353 were excluded (based on exclusion criteria), resulting in 41228 enrolments. Among these, 130 had no test results, and 27 had inconclusive results, resulting in 41071 participants with either RT-PCR or IgG antibody or both test results available. Further, 40030 had valid IgG test outcomes, while 1041 had invalid, or inconclusive, or unavailable IgG test outcomes. Similarly, 39779 had valid RT-PCR test outcomes, and 1292 had invalid, inconclusive, or unavailable RT-PCR test outcomes (Supplementary Figure 3).

#### IgG prevalence

Assuming the laboratory-calibrated 92.2% *analytical sensitivity* and 97.7% specificity for the ELISA-kit, the overall weighted adjusted seroprevalence of IgG in Round 2 was 15.6% (95% CI: 14.9–16.3), as of 18 February 2021, which is the end date for Round 2 (Table 1). Based on the 6002 positive and 34028 negative outcomes, among the 40030 valid IgG outcomes, the crude IgG prevalence was 6002/40030 = 15.0%.

**Table 1:** Seroprevalence of IgG antibodies against SARS-CoV2 and Active Infection in Karnataka at the end of Round 2.

| Category |  | Type | Samples<br>y | %-IgG against SARS-CoV2® | %-Active Infection<br>of COVID-19® | %-Prevalence of<br>COVID-19® | Odds Ratio |
| --- | --- | --- | --- | --- | --- | --- | --- |
| State | Karnataka | Crude | 41071 | 6002/40030 | 187/39779 | 6161/41071 | - |
|  |  | Adjusted | 41228 | 15.5 | 0 | 15.5 |  |
|  |  | Weighted Adjusted | 41228 | 15.6 (14.9--16.3) | 0 (0--0.3) | 15.6 (14.8--16.4) |  |
| Demography | Sex | Male | 19165 | 15.4 (14.4--16.4) | 0 (0--0.5) | 15.4 (14.3--16.5) | 1.22 (1.03--1.45) |
|  |  | Female | 22046 | 13 (12.1--13.9) | 0 (0--0.4) | 13 (12--13.9) | 1 |
|  |  | Other | 17 | 36.7 (0--80.6) | 0 (0--15.7) | 36.7 (0--82.5) | 3.88 (0--34.57) |
|  | Age | 18 - 29 | 15841 | 10.8 (9.8--11.7) | 0 (0--0.5) | 10.8 (9.7--11.9) | 1 |
|  |  | 30 - 39 | 7856 | 14.1 (12.5--15.6) | 0 (0--0.7) | 14.1 (12.4--15.7) | 1.36 (1.05--1.73) |
|  |  | 40 - 49 | 5745 | 17.4 (15.5--19.4) | 0 (0--0.8) | 17.4 (15.3--19.5) | 1.74 (1.34--2.26) |
|  |  | 50 - 59 | 3967 | 16.8 (14.5--19.2) | 0 (0--1) | 16.8 (14.3--19.3) | 1.67 (1.24--2.23) |
|  |  | 60 and above | 7818 | 17.3 (15.6--18.9) | 0 (0--0.7) | 17.3 (15.5--19.1) | 1.73 (1.36--2.2) |
|  | Region | Rural | 4074 | 15.4 (13.2--17.6) | 0 (0--1) | 15.4 (13--17.8) | 1 |
|  |  | Urban | 37154 | 14 (13.3--14.7) | 0 (0--0.3) | 14 (13.2--14.8) | 0.89 (0.7--1.16) |
| Risk Category |  | High-risk<sup>#</sup> | 13865 | 16.8 (15.6--18) | 0 (0--0.5) | 16.8 (15.5--18.1) | 1.6 (1.3--1.99) |
|  |  | Moderate-risk | 13714 | 14.3 (13.2--15.5) | 0 (0--0.5) | 14.3 (13.1--15.6) | 1.32 (1.06--1.66) |
|  |  | Low-risk | 13649 | 11.2 (10.1--12.3) | 0 (0--0.5) | 11.2 (10--12.4) | 1 |
| Risk Sub-category | High-risk | Elderly | 6740 | 17.3 (15.5--19.1) | 0 (0--0.8) | 17.3 (15.4--19.2) | 2.14 (1.55--3.02) |
|  |  | Persons with comorbidities | 7125 | 16.3 (14.6--18) | 0 (0--0.8) | 16.3 (14.5--18.2) | 1.99 (1.45--2.83) |
|  | Moderate-risk | Bus conductors/Auto drivers | 2694 | 16.5 (13.7--19.3) | 0 (0--1.2) | 16.5 (13.5--19.5) | 2.02 (1.33--3.08) |
|  |  | Pourakarmikas / waste collectors | 2665 | 14.8 (12.1--17.5) | 0 (0--1.2) | 14.8 (11.8--17.7) | 1.78 (1.14--2.73) |
|  |  | Healthcare workers | 2701 | 15 (12.3--17.7) | 0 (0--1.2) | 15 (12.1--17.9) | 1.81 (1.17--2.77) |
|  |  | Vendors at vegetable markets | 2715 | 13.3 (10.8--15.9) | 0 (0--1.2) | 13.3 (10.5--16.2) | 1.57 (1--2.45) |
|  |  | Congregate settings<sup>§</sup> | 2939 | 12.3 (9.9--14.7) | 0 (0--1.2) | 12.3 (9.6--14.9) | 1.44 (0.91--2.22) |
|  | Low-risk | Outpatient department | 6876 | 13.5 (11.9--15.1) | 0 (0--0.8) | 13.5 (11.7--15.3) | 1.6 (1.13--2.29) |
|  |  | Pregnant women | 6773 | 8.9 (7.5--10.3) | 0 (0--0.8) | 8.9 (7.3--10.5) | 1 |
| Pre-existing medical conditions |  | More than one | 1067 | 19.1 (14.5--23.8) | 0 (0--2) | 19.1 (14.2--24.1) | 1.46 (0.97--2.11) |
|  |  | One | 4808 | 15.1 (13.1--17.1) | 0 (0--0.9) | 15.1 (12.9--17.3) | 1.1 (0.87--1.39) |
|  |  | None | 35353 | 13.9 (13.1--14.6) | 0 (0--0.3) | 13.9 (13.1--14.6) | 1 |
| Symptoms |  | More than one | 1037 | 15.3 (10.9--19.6) | 0 (0--2) | 15.3 (10.5--20) | 1.07 (0.65--1.59) |
|  |  | One | 6026 | 12.6 (10.9--14.3) | 0 (0--0.8) | 12.6 (10.7--14.5) | 0.86 (0.67--1.08) |
|  |  | None | 34165 | 14.4 (13.6--15.1) | 0 (0--0.3) | 14.4 (13.6--15.2) | 1 |

<sup>‡</sup> Includes only samples that have been mapped to participants.
<sup>®</sup> All estimates are adjusted for sensitivities and specificities of the RT-PCR and antibody testing kits and procedures; the assumed values are RT-PCR sensitivity 0.95, specificity 0.97, IgG ELISA kit sensitivity 0.921, specificity 0.977; Weighted estimates for Karnataka estimate the prevalence in each unit and then weights according to population
<sup>§</sup> Markets, Malls, Retail stores, Bus stops, Railway stations, waste collectors; <sup>#</sup>Some individuals recruited in the moderate and low-risk categories, but with high risk-features, were moved to high-risk.

#### Active infection

The weighted adjusted active infection was estimated to be 0.0% (95% CI: 0.0–0.3) during the Round 2 period. Based on the 187 positive and 39592 negative outcomes, among the 40030 valid RT-PCR outcomes, the crude active prevalence was 187/39779 = 0.5% (Table 1).

#### Total prevalence

We estimated the overall weighted adjusted seroprevalence as 15.6% (95% CI: 14.8–16.4) (Table 1).

#### Demography

The total prevalence among males and females was 15.4% (14.3–16.5) and 13.0% (12.0--13.9), respectively. The total prevalence among 18-29, 30-39, 40-49, 50-59, and 60+ age-groups were 10.8% (9.7--11.9), 14.1% (12.5--15.7), 17.4% (15.3--19.5), 16.8% (14.3--19.3), and 17.3% (15.5--19.1), respectively. Thus, the total prevalence was higher among males than females and was higher among the elderly population when compared with those aged <30 years (Table 1).

#### Stratifications

The high-risk (vulnerable) segment of the population continued to be at higher risk (16.8% (15.5--18.1)), followed by the moderate risk (14.3% (13.1--15.6)), and then the low-risk population (11.2% (10.0--12.4)). In a reversal from Round 1, the rural population had a higher total prevalence (15.4% (13.0--17.8)) compared to the urban population (14% (13.2-- 14.8)); this is unadjusted for the excluded population due to lack of availability of fine-grained rural/urban case data (Table 1).

Across risk-subcategories, pregnant women had the least total prevalence (8.9% (7.3--10.5)), while bus-conductors/auto-drivers (16.5% (13.5--19.5)), people with co-morbidities (16.3% (14.5--18.2)), and the elderly (17.3% (15.4--19.2)) had higher prevalence. Interestingly, *pourakarmikas*, who carry out work in less hygienic conditions, had a total prevalence of 14.8% (11.8--17.7) that did not stand out from the general population.

#### Odds risk for detectable IgG antibodies

The odds for males was 1.22 as compared to females. Across age groups, the odds for the 30-39, 40-49, 50-59, and 60+ age groups, over the reference 18-29 age group, were 1.36, 1.74, 1.67, and 1.73, respectively. The vulnerable population in the high-risk category continued to have higher odds of 1.6 over the low-risk category. In contrast, the moderate-risk category had odds of 1.32 over the low-risk category. The elderly had higher odds of 2.14 over the reference pregnant women sub-category. The odds for the urban population were 0.89 as compared to the rural population. See Table 1 for confidence intervals.

#### Pre-existing medical conditions

The seroprevalence of IgG antibodies was higher among those with more than one co-morbidity (19.1%), followed by those with one co-morbidity (15.1%). Those who reported having more than one symptom had a higher IgG prevalence (15.3%) than those with no symptoms (14.4%).

#### Cases-to-infections ratio (CIR)

At the state level, for every RT-PCR confirmed case, there were 12 infected individuals with detectable IgG levels (Table 2). This was estimated using the 946860 reported cases in Karnataka as of 18 February 2021. The CIR across units ranged from 3 (Rest of Bengaluru Urban) to 39 (Belgaum), with the CIR of Bengaluru Urban Conglomerate as 6.

**Table 2:** Seroprevalence of IgG antibodies against SARS-CoV2 and Active Infection in districts of Karnataka state at the end of Round 2 (n=41228)

| Unit | Samples<sup>y</sup> | %-IgG against SARS-CoV2<sup>@</sup> | %-Active Infection of COVID-19<sup>@</sup> | %-Prevalence of COVID-19<sup>@</sup> |
| --- | --- | --- | --- | --- |
| Karnataka | 41228 | 15.6 (14.9--16.3) | 0 (0--0.3) | 15.6 (14.8--16.4) |
| Mysuru | 1104 | 33.6 (28.2--39) | 0 (0--1.9) | 33.6 (28--39.3) |
| Mandya | 1159 | 31.9 (26.9--37) | 0 (0--1.8) | 31.9 (26.6--37.3) |
| Kodagu | 1063 | 27.1 (22.1--32.1) | 0 (0--1.9) | 27.1 (21.8--32.4) |
| Chamarajanagar | 1161 | 22.6 (17.6--27.6) | 0 (0--1.9) | 22.6 (17.3--27.9) |
| Kolar | 1050 | 20.8 (16.1--25.4) | 0 (0--1.9) | 20.8 (15.8--25.8) |
| Bengaluru Rural | 1084 | 20.3 (15.7--24.8) | 0 (0--2) | 20.3 (15.4--25.1) |
| Dakshina Kannada | 1074 | 19.8 (15.4--24.3) | 0 (0--1.9) | 19.8 (15.1--24.6) |
| Belgaum | 1110 | 19.4 (14.9--23.9) | 0 (0--1.9) | 19.4 (14.5--24.2) |
| Bengaluru Urban Conglomerate | 9730 | 18.7 (17.1--20.2) | 0 (0--0.7) | 18.7 (17--20.4) |
| Udupi | 1076 | 17.9 (13.7--22.1) | 0 (0--1.9) | 17.9 (13.4--22.5) |
| Chitradurga | 1060 | 16.6 (12.3--21) | 0 (0--1.9) | 16.6 (11.9--21.3) |
| Davanagere | 1054 | 16.2 (11.9--20.4) | 0 (0--2) | 16.2 (11.6--20.8) |
| Bagalkot | 1051 | 15.7 (11.5--19.9) | 0 (0--1.9) | 15.7 (11.1--20.3) |
| Ramanagar | 1057 | 14.5 (10.5--18.6) | 0 (0--1.9) | 14.5 (10.1--19) |
| Chikkaballapur | 1062 | 13.7 (9.7--17.7) | 0 (0--1.9) | 13.7 (9.3--18.1) |
| Gadag | 1137 | 13.1 (9.4--16.9) | 0 (0--1.9) | 13.1 (9--17.3) |
| Vijayapura | 1058 | 12.9 (9--16.8) | 0 (0--1.9) | 12.9 (8.6--17.3) |
| Shivamogga | 1062 | 12.8 (8.9--16.6) | 0 (0--1.9) | 12.8 (8.5--17) |
| Chikmagalur | 1050 | 12.6 (8.8--16.4) | 0 (0--1.9) | 12.6 (8.4--16.8) |
| Ballari | 1056 | 12.3 (8.5--16) | 0 (0--1.9) | 12.3 (8.1--16.5) |
| Tumakuru | 1051 | 10.7 (7.1--14.4) | 0 (0--2) | 10.7 (6.6--14.9) |
| Raichur | 1247 | 10.5 (7.1--13.9) | 0 (0--1.8) | 10.5 (6.7--14.3) |
| Uttara Kannada | 1080 | 10.3 (6.7--13.8) | 0 (0--1.9) | 10.3 (6.3--14.3) |
| Koppal | 1063 | 9 (5.6--12.4) | 0 (0--1.9) | 9 (5.2--12.8) |
| Hassan | 1051 | 7.6 (4.6--10.6) | 0 (0--2) | 7.6 (4--11.2) |
| Kalaburagi | 1087 | 6.3 (3.3--9.2) | 0 (0--1.9) | 6.3 (2.8--9.8) |
| Dharwad | 1101 | 5.8 (3--8.5) | 0 (0--1.9) | 5.8 (2.4--9.1) |
| Yadgir | 1061 | 5.5 (2.7--8.4) | 0 (0--1.9) | 5.5 (2.1--9) |
| Bidar | 1168 | 4.5 (1.9--7.1) | 0 (0--1.9) | 4.5 (1.3--7.7) |
| Haveri | 1061 | 3.7 (1.2--6.1) | 0 (0--1.9) | 3.7 (0.5--6.8) |

<sup>y</sup> Includes only samples that have been mapped to individuals.
<sup>@</sup> Adjusted for sensitivities and specificities of RT-PCR, and antibody testing kits and procedures.

#### Infection fatality rate (IFR)

The IFR was estimated to be 0.11% statewide and ranged from 0.02% (Chitradurga) to 0.50% (Dharwad), with 19 out of 38 units below the state IFR. As in Round 1, the Dharwad district had the highest IFR (Table 2). The IFR of Bengaluru Urban Conglomerate was 0.17%.

#### Districts/unit variations across the state

The active infection fractions across all districts were estimated as 0.0% (with varying confidence intervals given in Table 3). Hence, the total prevalence is the same as the IgG prevalence, with minor expansions of the confidence intervals. The total prevalence was highest in Mysuru district (33.6% (28.0--39.3)), followed by Mandya (31.9% (26.6--37.3)), Kodagu (27.1% (21.8--32.4)), Chamarajanagar (22.6% (17.3--27.9)), and Kolar (20.8% (15.8--25.8)). Other units reported ≥15% seroprevalence were Bengaluru Rural, Dakshina Kannada, Belgaum, Bengaluru Urban Conglomerate (18.7% (17-- 20.4)), Udupi, Chitradurga, Davanagere and Bagalkot. Haveri district had the lowest seroprevalence (3.7% (0.5--6.8)).

**Table 3:**
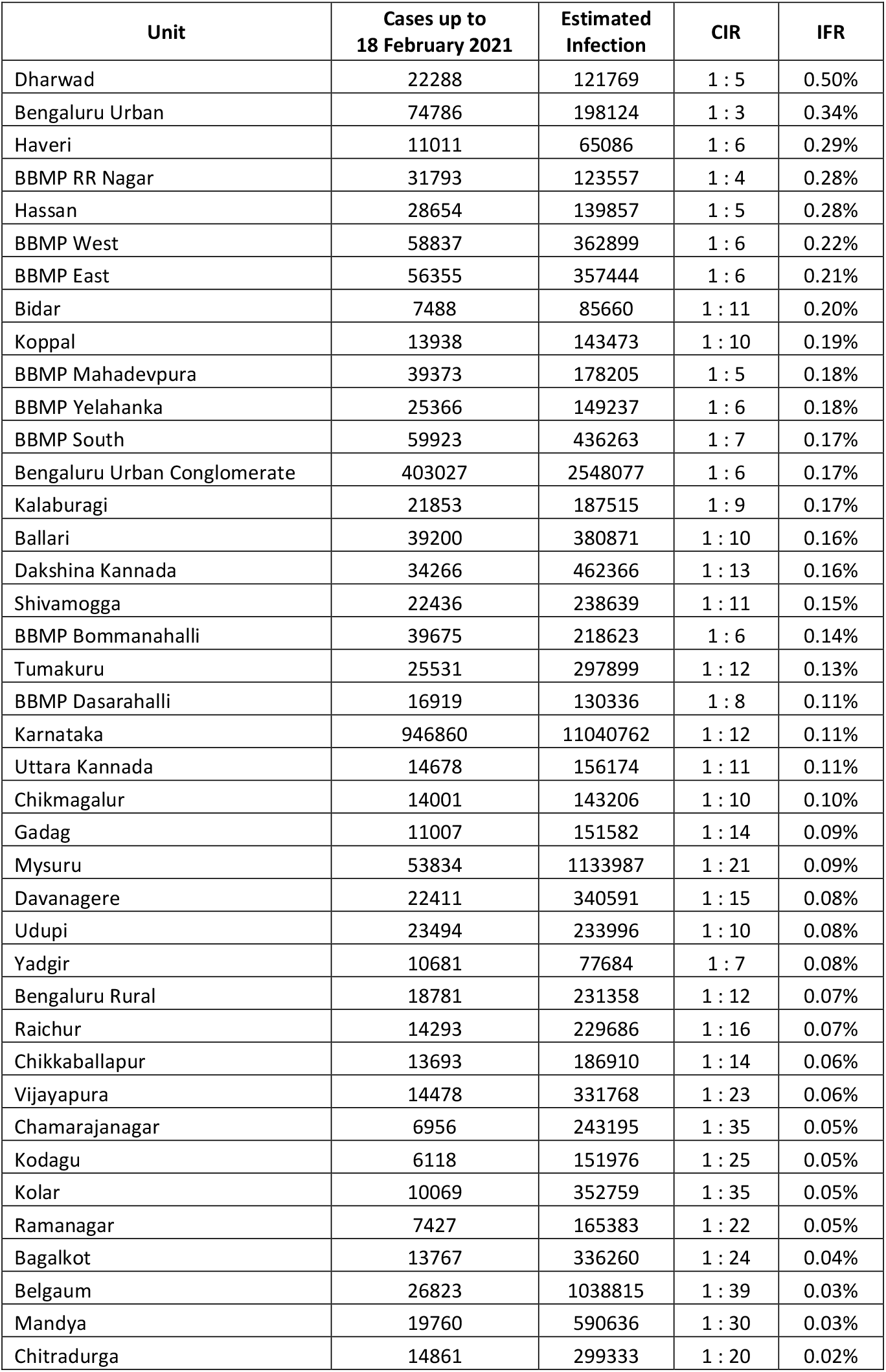
**CIR and IFR across all 30 districts in Karnataka. Note that the CIR estimate is likely to be conservative and the IFR pessimistic on account of the low sensitivity of the kit for a population with infection in the past**.

| Unit | Cases up to<br>18 February 2021 | Estimated<br>Infection | CIR | IFR |
| --- | --- | --- | --- | --- |
| Dharwad | 22288 | 121769 | 1 : 5 | 0.50% |
| Bengaluru Urban | 74786 | 198124 | 1 : 3 | 0.34% |
| Haveri | 11011 | 65086 | 1 : 6 | 0.29% |
| BBMP RR Nagar | 31793 | 123557 | 1 : 4 | 0.28% |
| Hassan | 28654 | 139857 | 1 : 5 | 0.28% |
| BBMP West | 58837 | 362899 | 1 : 6 | 0.22% |
| BBMP East | 56355 | 357444 | 1 : 6 | 0.21% |
| Bidar | 7488 | 85660 | 1 : 11 | 0.20% |
| Koppal | 13938 | 143473 | 1 : 10 | 0.19% |
| BBMP Mahadevpura | 39373 | 178205 | 1 : 5 | 0.18% |
| BBMP Yelahanka | 25366 | 149237 | 1 : 6 | 0.18% |
| BBMP South | 59923 | 436263 | 1 : 7 | 0.17% |
| Bengaluru Urban Conglomerate | 403027 | 2548077 | 1 : 6 | 0.17% |
| Kalaburagi | 21853 | 187515 | 1 : 9 | 0.17% |
| Ballari | 39200 | 380871 | 1 : 10 | 0.16% |
| Dakshina Kannada | 34266 | 462366 | 1 : 13 | 0.16% |
| Shivamogga | 22436 | 238639 | 1 : 11 | 0.15% |
| BBMP Bommanahalli | 39675 | 218623 | 1 : 6 | 0.14% |
| Tumakuru | 25531 | 297899 | 1 : 12 | 0.13% |
| BBMP Dasarahalli | 16919 | 130336 | 1 : 8 | 0.11% |
| Karnataka | 946860 | 11040762 | 1 : 12 | 0.11% |
| Uttara Kannada | 14678 | 156174 | 1 : 11 | 0.11% |
| Chikmagalur | 14001 | 143206 | 1 : 10 | 0.10% |
| Gadag | 11007 | 151582 | 1 : 14 | 0.09% |
| Mysuru | 53834 | 1133987 | 1 : 21 | 0.09% |
| Davanagere | 22411 | 340591 | 1 : 15 | 0.08% |
| Udupi | 23494 | 233996 | 1 : 10 | 0.08% |
| Yadgir | 10681 | 77684 | 1 : 7 | 0.08% |
| Bengaluru Rural | 18781 | 231358 | 1 : 12 | 0.07% |
| Raichur | 14293 | 229686 | 1 : 16 | 0.07% |
| Chikkaballapur | 13693 | 186910 | 1 : 14 | 0.06% |
| Vijayapura | 14478 | 331768 | 1 : 23 | 0.06% |
| Chamarajanagar | 6956 | 243195 | 1 : 35 | 0.05% |
| Kodagu | 6118 | 151976 | 1 : 25 | 0.05% |
| Kolar | 10069 | 352759 | 1 : 35 | 0.05% |
| Ramanagar | 7427 | 165383 | 1 : 22 | 0.05% |
| Bagalkot | 13767 | 336260 | 1 : 24 | 0.04% |
| Belgaum | 26823 | 1038815 | 1 : 39 | 0.03% |
| Mandya | 19760 | 590636 | 1 : 30 | 0.03% |
| Chitradurga | 14861 | 299333 | 1 : 20 | 0.02% |

#### Bengaluru metropolitan area

Within the Bengaluru metropolitan area (Bruhat Bengaluru Mahanagara Palike (BBMP)), the total prevalence varied from 13.8% (BBMP RR Nagara) to 24.3% (BBMP Dasarahalli) (Supplementary Table 1). The CIR ranged from 4–8 and the IFR from 0.11%--0.28% (Supplementary Table 1).

#### Explanatory variables

Logistic regression indicated that the following factors led to a higher probability of a positive IgG test outcome: “Other” sex category, chronic renal disease, moderate- or high-risk category, attenders of outpatients, transport professionals (bus-conductors/auto-drivers), healthcare workers, and age 30 years and above (Supplementary Figure 4, Table 4). No significant association was observed between symptoms and the presence of IgG antibodies.

**Table 4:** Logistic regression for predicting IgG prevalence.

| Features | $\beta_L$ | $\sigma_L$ | Logistic p-val |
| --- | --- | --- | --- |
| Intercept | -2.2 | 0.06 | *** |
| Chronic Renal Disease | 0.63 | 0.3 | * |
| Moderate Risk | 0.21 | 0.074 | ** |
| High Risk | 0.3 | 0.071 | *** |
| OPD attendee | 0.27 | 0.057 | *** |
| Bus conductors, Auto drivers | 0.2 | 0.077 | ** |
| Age 30-39 years | 0.17 | 0.043 | *** |
| Age 40-49 years | 0.36 | 0.048 | *** |
| Age 50-59 years | 0.32 | 0.057 | *** |
| Age 60+ years | 0.34 | 0.079 | *** |
| Sex: Other | 1.2 | 0.51 | * |
| Region: Urban | -0.14 | 0.046 | ** |
| Urbanisation | 0.28 | 0.056 | *** |

\*\*\* indicates a p-value of less than 0.001, \*\* indicates less than 0.01, \* indicates less than 0.05.

### Longitudinal study for estimating the clinical sensitivity of the IgG ELISA kit

The longitudinal study was done from 02 April to 11 May 2021. We collected 648 samples (after removing one duplicate) from 26 units, yielding a participation rate of 648/4420 = 14.7%. The units that did not have participants were Gadag, Raichur, Kalaburagi, Dharwad, BBMP South, BBMP East, BBMP West, BBMP RR Nagar, BBMP Mahadevpura and BBMP Yelahanka.

Out of the 648 samples, only 370 IgG ELISA test outcomes were valid based on controls. Of these, 144 tested positive and 226 tested negative. Thus, only 38.9% of the first-round positive participants were above the detection threshold of the IgG ELISA test kit during the time frame of the longitudinal study. Given the significant lapse of time between the end of Round 2 and the median time of the longitudinal study (22 April 2021), we deduce that the clinical sensitivity of the IgG ELISA test kit is ≥38.9% at the time of Round 2.

#### Upper bound on the total disease burden based on the longitudinal study

Assuming a clinical sensitivity ≥38.9%, following the same statistical analysis, the total number infected in Karnataka as of 18 February 2021 was ≤35.8% (95% CI: 34.0—37.7), CIR ≤27, and IFR ≥0.05%. Given the total burden of 27.7% (95% CI: 26.1–29.3), measured at the end of Round 1,[17] we conclude that the COVID-19 burden of Karnataka was between 26.1–37.7% (at 90% confidence) with CIR range 12—27 and IFR range 0.24%--0.50%, as of 18 February 2021. Dharwad’s IFR, the highest, ranged from 0.24%--0.50%.

## DISCUSSION

Similar to the first round, our present study involves several district and state agencies: 290 healthcare facilities across all districts of Karnataka and the associated healthcare workers participated in the effort. Our study is further unique in jointly estimating active prevalence and IgG antibody prevalence. Despite the sentinel-based nature of the survey, our sampling frame attempted to overcome bias in the facility-based sampling frame by, for example, sampling from pregnant women coming for a regular check-up and sampling attendees of patients instead of the patients themselves [17]. Additionally, to account for IgG antibody waning, we conducted a longitudinal study for estimating the clinical sensitivity of the IgG ELISA kit, and this enabled an interval estimate of the total prevalence in the state.

The estimated IgG prevalence at the end of Round 2 (15.6%) is remarkably lower than the estimated total infection of 27.7% (95% CI: 26.1–29.3) at the end of Round 1 (IgG prevalence 16.8% (15.5–18.1)).[17] Tamil Nadu, a neighbouring state, also reported a reduction in March-April 2021 (23%) compared to October-November 2020 (31.6%).[35] Assuming the lab-calibrated analytical sensitivity (92.2%) yields an under-estimate of the IgG prevalence in view of IgG level decline (Round 2 began 131 days after Round 1 and 98 days after the active cases peaked in the state). The ICMR third round study<u>[22]</u> took two approaches to handle antibody waning – reduction in the optical density thresholds and an independent validation of the testing kit – and reported the adjustments. We conducted an independent validation via a longitudinal study.

The longitudinal study (conducted on a subset of the recalled Round 1 positive population) yielded a clinical sensitivity of ≥38.9% during the Round 2 period. The IgG ELISA test used the whole-cell antigen instead of the more specific recombinant nucleocapsid or spike protein antigens [25]. This, along with antibody waning, may have played a role in its reduced clinical sensitivity.

Given the lapse of time between Round 2 and the longitudinal study, the measured clinical sensitivity of 38.9% may be viewed as a lower bound on this sensitivity since fewer days would have elapsed between the date of infection of positive participants and the end of Round 2. By assuming this pessimistic 38.9% value of clinical sensitivity, following the same statistical analysis, we estimated that *at most*, 35.8% (95% CI: 34.0-37.7) were infected in Karnataka, as of 18 February 2021. Together with the total burden of 27.7% (95% CI: 26.1–29.3), estimated at the end of Round 1,[17], we concluded that Karnataka’s COVID-19 burden was between 26.1–37.7% (at 90% confidence), suggesting a significant level of susceptibility (and hence insufficient natural immunity) in the population as of 18 February 2021.

The estimated active prevalence was 0.0% across all districts. The subsequent rise in infection from March to June 2021 may be due to a combination of effects ranging from immunity waning[23] to the emergence of the B.1.617 variant and its sub-variants.[37]

Comparison of the CIR range 12--27 and IFR range 0.05--0.11% (Round 2) with CIR 40 and IFR 0.05% (Round 1) for Karnataka suggests improved case identification between Round 1 and Round 2.

As in Round 1, Dharwad had the highest IFR (0.24%–0.50%). This could be due to reporting differences or issues related to clinical practice or travel from neighbouring units to avail critical or tertiary health care facilities at Dharwad.[17] Further research should explore these hypotheses.

Males continued to be at higher risk than females (odds ratio 1.22), the vulnerable population in the high-risk category continued to be at higher risk than the low-risk category (odds ratio 1.6), those in the higher age groups continued to be at higher risk than the 18-29 age group (Table 1). However, rural areas were more at risk than urban areas (odds ratio urban 0.89 < 1 rural), a reversal from Round 1. Together with the observations on antibody waning, the higher risk for rural areas suggests that the infection continued to be active in the rural areas after it had subsided in the urban areas during October 2020 – February 2021.

Pregnant women are known to be more susceptible to respiratory pathogens, and hence to SARS-CoV-2, than the general population [36]. It is, therefore, reassuring to note that the total prevalence among pregnant women was the lowest, suggesting the hypothesis that their behavioural patterns result in significantly lower contact rates.

Serial serosurveys repeated at the same sites can enable the comparison of epidemiological metrics across time. A comparison of IgG prevalence alone between Round 1 and Round 2 suggests that about 17 units have lower IgG prevalence in Round 2, while the remaining 21 have higher IgG prevalence (Figure 1). However, when we compare the total prevalence of Round 1 and Round 2, the latter is mostly lower except for a marginal increase in 11 units (Supplementary Figure 5), possibly due to the reduced clinical sensitivity of the IgG ELISA test kit. Another interesting observation is that while high urbanisation leads to lower CIR (Figure 2), some districts with low urbanisation have low CIR. However, some others have higher CIR, suggesting the need to step up surveillance in those latter rural units (Belgaum, Kolar, Chamarajanagar, and Mandya). Finally, as in [17], Figure 3 suggests a possible classification of districts into those with high/low CIR and high/low IFR. Districts with high CIR and low IFR in the top-left quadrant should consider re-evaluating their testing strategies and death reporting.

**Figure 1:**
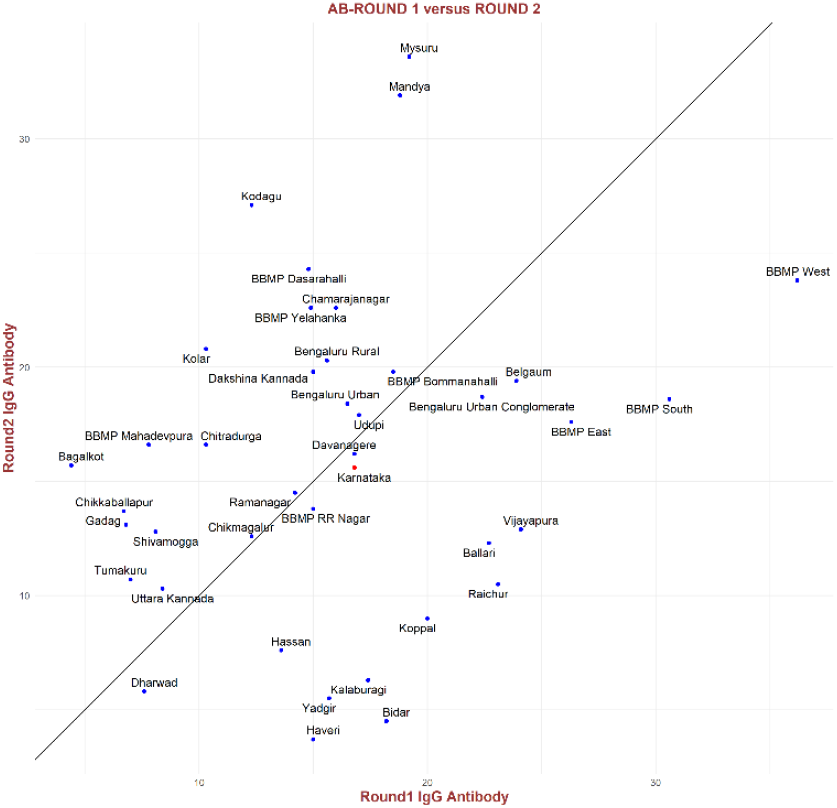
Comparing IgG prevalence across Round 1 and Round 2, IgG increased in about 21/38 units (above the line) while it decreased in 17/38 units (below the line).

**Figure 2:**
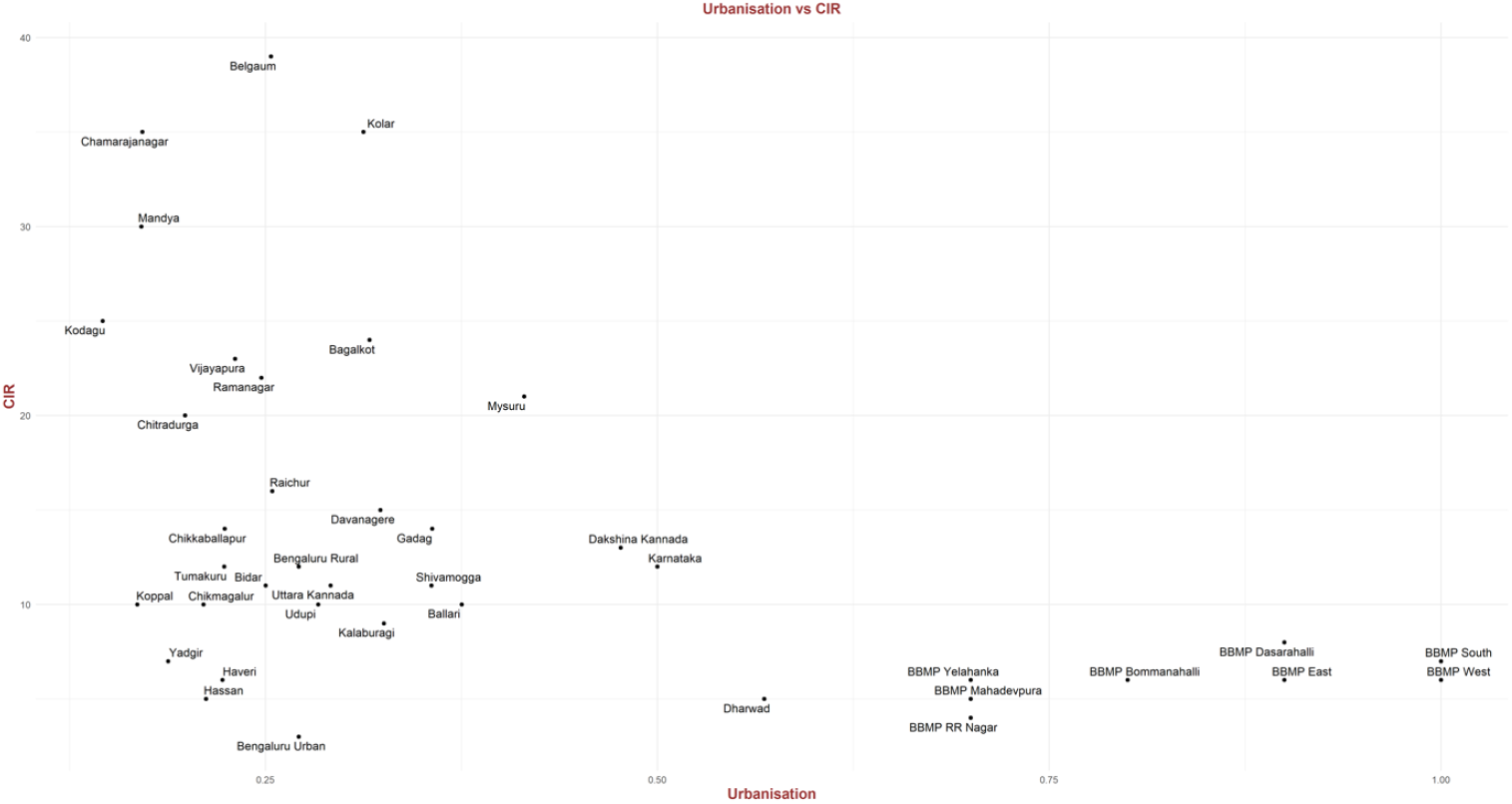
CIR as a function of urbanisation. Observe that the higher the urbanisation value, the lower the CIR. Some locations with lesser urbanisation also have lower CIR. However, some others have higher CIR, suggesting that these units are missing regions of circulation of the virus and could benefit from increased surveillance.

**Figure 3:**
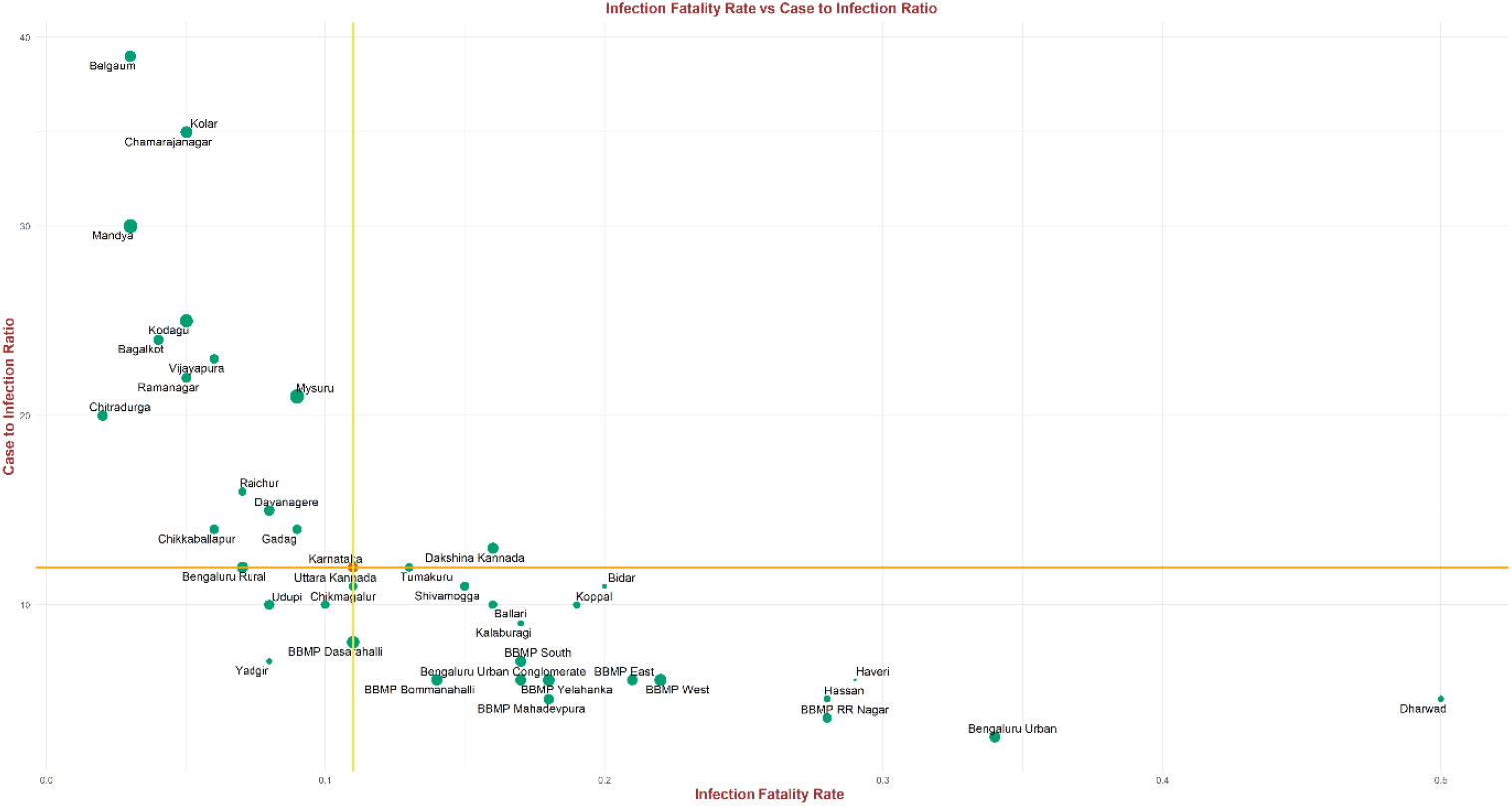
The IFR versus CIR in the districts of Karnataka. Districts in the top-left quadrant, with low IFR and high CIR, may have to re-evaluate both their testing strategies and death reporting.

As highlighted above, the sentinel-based population survey strategy has enabled the identification of trends over time. Such a survey is also easier to implement in terms of planning logistics for quick deployment. The study findings enable identifying districts that need better testing, reporting, or clinical management, all of which ultimately reduce the number of deaths due to COVID-19. Since the state was far from attaining natural immunity, vaccination coverage should be stepped up.

## Data Availability

The data are accessible to researchers upon formal request for data addressed to the Commissioner, Health and Family Welfare Services, Government of Karnataka.

## CONTRIBUTORS

The survey was a collaborative effort of the Department of Health and Family Welfare, National Institute of Mental Health and Neuro-Sciences, Indian Institute of Public Health - Bengaluru, Indian Institute of Science, Indian Statistical Institute (Bengaluru Centre), UNICEF, MS Ramaiah Medical College, Bangalore Medical College, and others. Giridhara R Babu, R Lalitha Hande, M Rajagopal Padma, Siva Athreya and Rajesh Sundaresan designed the protocol for the second round. Mysore Kalappa Sudarshan and Anita Desai reviewed and provided feedback on the design, the implementation of the survey, the analyses, and helped articulate the findings. Jawaid Akhtar and Trilok Chandra reviewed the protocol, led the implementation of the survey, and identified district-level public health responses. M Rajagopal Padma and Parimala S. Maroor coordinated the implementation across the state and reviewed the manuscript. Deepa Ravi, Shilpa Shiju and Prameela Dinesh developed the detailed protocol and standard operating procedures (protocol manual), implemented the study, and reviewed the manuscript. Siva Athreya and Rajesh Sundaresan planned and executed the data analysis, arrived at the initial findings. Deepa Ravi, Eunice Lobo, Yamuna Ana, and Prafulla Shriyan drafted the manuscript. Ambica Rangaiah, Ashok Munivenkatappa, Krishna S, Shantala Gowdara Basawarajappa, HG Sreedhara, Siddesh KC, Amrutha Kumari B, Nawaz Umar, Mythri BA developed the lab protocols and provided the test results. Mythri KM contributed to IgG testing of sub study samples. Ravi Vasanthapuram designed and developed the protocol and revised the manuscript. All authors reviewed and approved the final manuscript.

## ACKNOWLEDGMENTS

We would like to express our thanks to Dr Arundathi, IAS, MD – NHM, and Dr. Om Prakash Patil Director – DHFWS, State Surveillance Unit, and Ms K Leelavathi, IAS, PD-KSAPS, for their support. We thank the DSOs, the DAPCU officers, the AMOs & Medical officers, the District Microbiologist and the District Epidemiologist and all other district-level staff for coordinating and implementing the survey, for guiding the health facility and laboratory staff in sample collection, and for coordinating sample transportation to mapped RT-PCR and antibody testing labs. We thank the District Surveillance teams and ICTC teams in the districts for the collection and transportation of COVID-19 samples. We thank the Lab Nodal Officers and staffs of ICMR labs for IgG antibody testing and RT-PCR testing. We thank Dr Kousalya R of Institute of Nephro Urology (INU) for contributing in IgG testing of Sub study samples. We thank Mr Ramesh, Mr Mahesh and Mr SreeRam from IT Cell Admin, E-Health Division for providing a web platform for metadata collection. We thank Dr Sathyam for help with data analysis and validation. Our heartfelt gratitude goes to all the lab technicians, counsellors – ICTC & NCDC, staff nurses, and health workers for filling data in the survey app, collecting samples, and sending them to the mapped laboratories. We also thank the Institute of Nephro Urology for some preliminary testing of samples using an alternative IgG testing kit. We thank all the study participants for providing their consent to be a part of this survey.

