## Supplementary Figure 1 for "Second round statewide survey for estimation of the burden of active infection and anti-SARS-CoV-2 IgG antibodies in the general population of Karnataka, India"

**1050**

**Low risk**

**350**

**Antibody**

**RTPCR**

**350**

**Moderate risk**

**Antibody**

**350**

**RTPCR**

**350**

**High risk**

**Antibody**

**350**

**RTPCR**

**350**

**Supplementary Figure 1: Schema for lab testing in each unit. Each unit was sampled from 1050 participants. They were divided equally among the three risk categories (low-risk, moderate-risk and high-risk categories). Nasopharyngeal/oropharyngeal swabs were taken for RT-PCR testing. Serum from 4 ml venous blood was extracted for IgG antibody testing.**
