## Supplementary Figure 2 for "Second round statewide survey for estimation of the burden of active infection and anti-SARS-CoV-2 IgG antibodies in the general population of Karnataka, India"

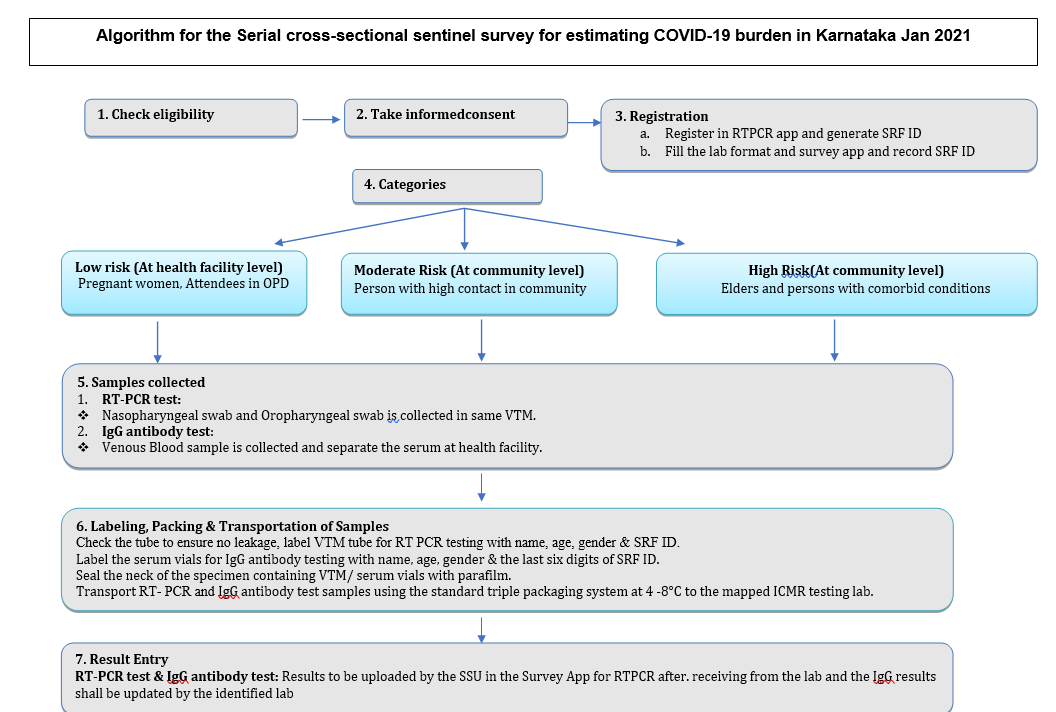

**Supplementary Figure 2: Algorithm for the serial cross-sectional sentinel survey for estimating COVID-19 burden in Karnataka.**
