## Supplementary Figure 3 for "Second round statewide survey for estimation of the burden of active infection and anti-SARS-CoV-2 IgG antibodies in the general population of Karnataka, India"

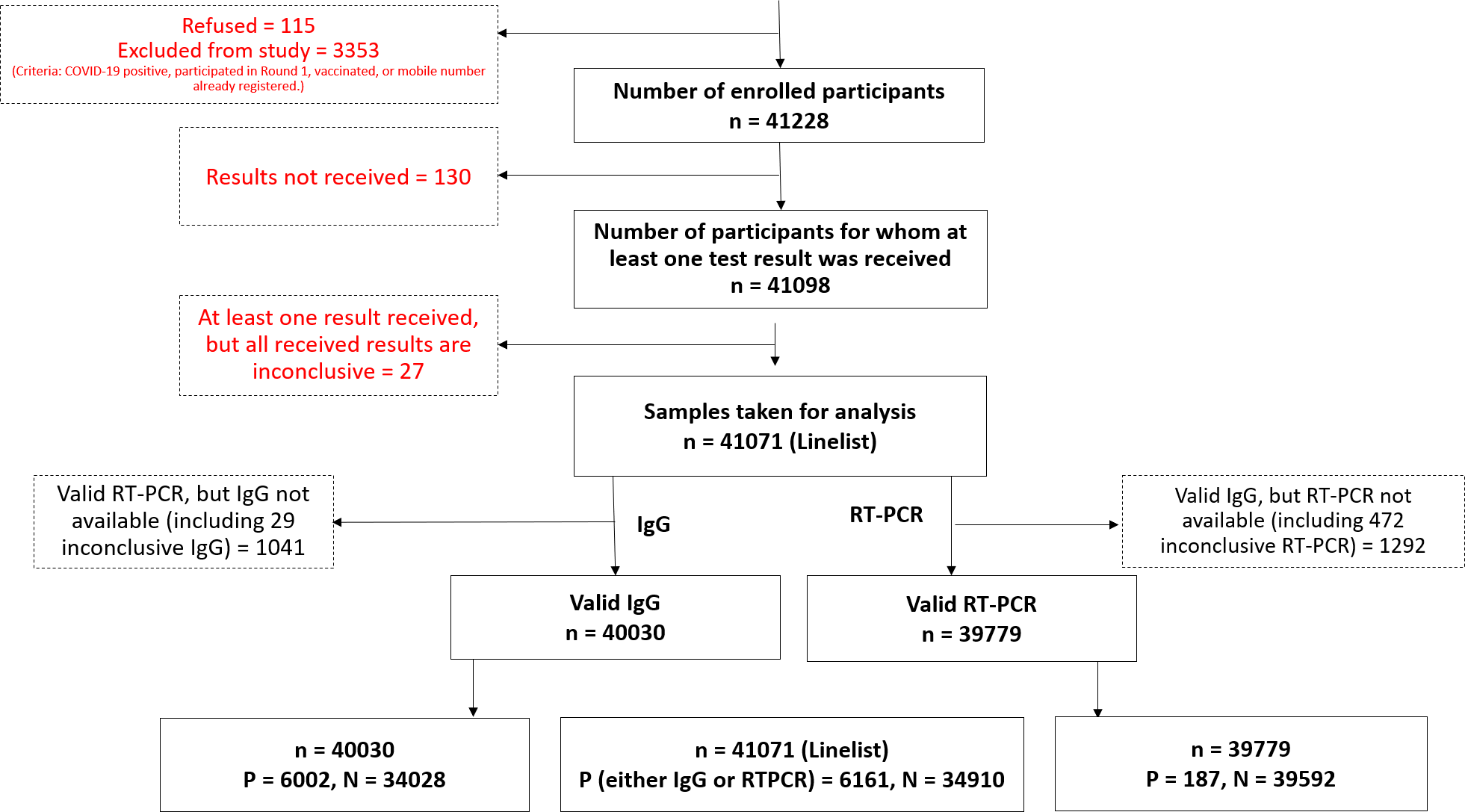

**Supplementary Figure 3: Flow chart of samples taken. Of the 44539 people who were approached, 115 refused participation and 3353 were excluded (based on exclusion criteria), resulting in the enrolment of 41228 participants in the different risk categories. Of these 41228 participants, 130 had no test results, and 27 had inconclusive results, resulting in 41071 participants with either RT-PCR or IgG antibody or both test results available. Among these 41071 participants, 40030 had valid IgG test outcomes, and 1041 had invalid, inconclusive or unavailable IgG test outcomes. Further, among these 41071 participants, 39779 had valid RT-PCR test outcomes, and 1292 had invalid, inconclusive, or unavailable RT-PCR test outcomes.**
