## Supplementary Figure 4 for "Second round statewide survey for estimation of the burden of active infection and anti-SARS-CoV-2 IgG antibodies in the general population of Karnataka, India"

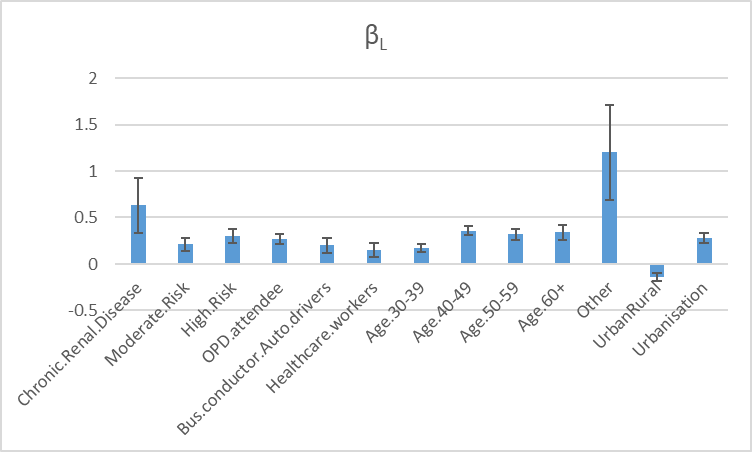

**Supplementary Figure 4: Weights on factors for predicting IgG positivity based on logistic regression. UrbanRural factor = Urban diminishes the chance of IgG positivity. All others increase the chance of IgG positivity, with those in the "Other" sex category and those with the chronic renal disease being at higher risk.**
