## Supplementary Figure 5 for "Second round statewide survey for estimation of the burden of active infection and anti-SARS-CoV-2 IgG antibodies in the general population of Karnataka, India"

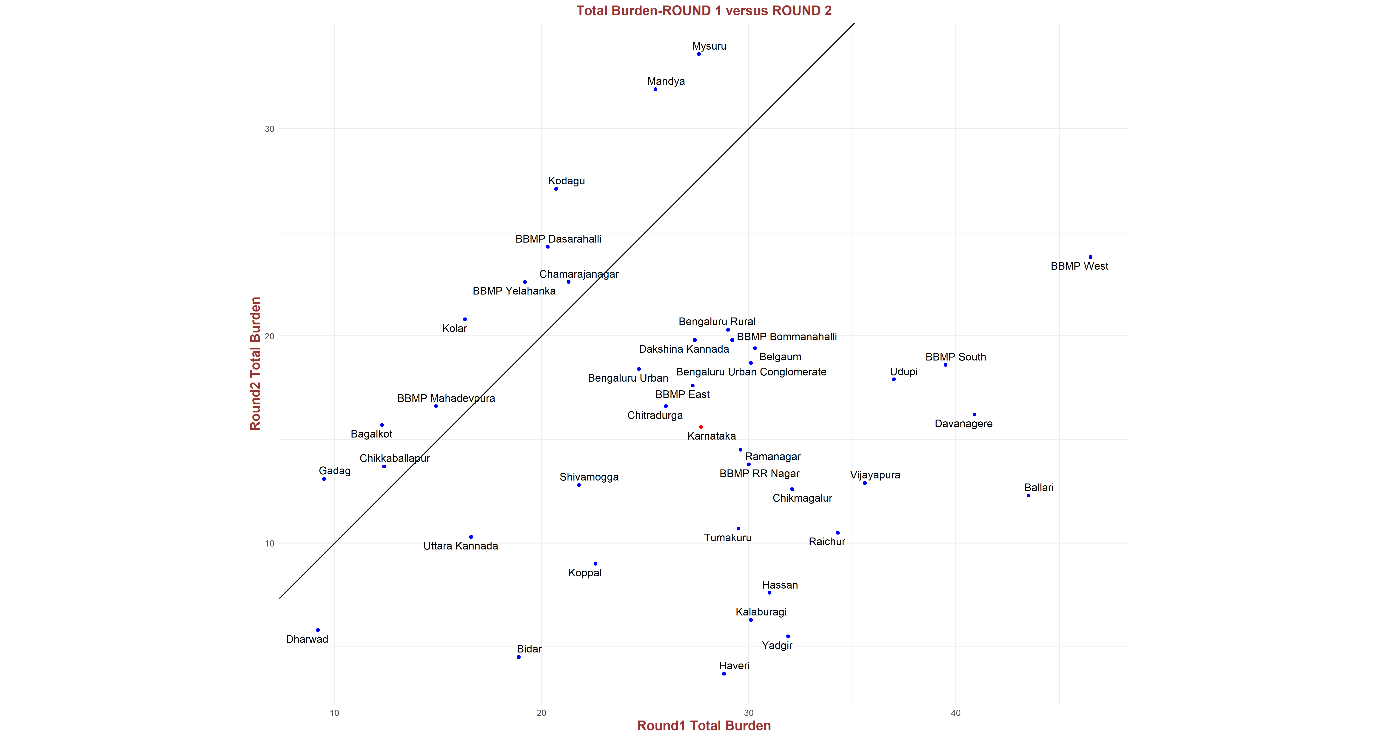

**Supplementary Figure 5: A comparison of the estimated total prevalence after Round 1 and Round 2 in the districts indicated lower total prevalence in 27/38 units suggesting a significant waning of IgG levels in the population.**
