## Supplementary Table 1 for "Second round statewide survey for estimation of the burden of active infection and anti-SARS-CoV-2 IgG antibodies in the general population of Karnataka, India"

**Supplementary Table 1: Seroprevalance of IgG antibodies against SARS-CoV-2 and active infection in Bengaluru Urban Conglomerate at the end of Round 2 (N = 9730).**

| **BBMP Zone** | **Samples^y^** | **%-IgG against**  **SARS-CoV2^@^** | **%-Active Infection of COVID-19^@^** | **%-Prevalence of COVID-19^@^** |
| --- | --- | --- | --- | --- |
| BBMP Dasarahalli | 1088 | 24.3 (19.6--29) | 0 (0--1.9) | 24.3 (19.3--29.3) |
| BBMP West | 1063 | 23.8 (18.8--28.7) | 0 (0--1.9) | 23.8 (18.5--29) |
| BBMP Yelahanka | 1112 | 22.6 (18.1--27) | 0 (0--2) | 22.6 (17.8--27.4) |
| BBMP Bommanahalli | 1070 | 19.8 (15.5--24.1) | 0 (0--1.9) | 19.8 (15.2--24.4) |
| **Bengaluru Urban Conglomerate** | **9730** | **18.7 (17.1--20.2)** | **0 (0--0.7)** | **18.7 (17--20.4)** |
| BBMP South | 1118 | 18.6 (14.5--22.8) | 0 (0--1.9) | 18.6 (14.1--23.2) |
| Bengaluru Urban | 1089 | 18.4 (14.8--22) | 0 (0--1.8) | 18.4 (14.4--22.4) |
| BBMP East | 1085 | 17.6 (13.5--21.7) | 0 (0--1.9) | 17.6 (13.1--22.1) |
| BBMP Mahadevpura | 1049 | 16.6 (12.5--20.6) | 0 (0--2) | 16.6 (12.1--21.1) |
| BBMP RR Nagar | 1056 | 13.8 (10.2--17.3) | 0 (0--1.9) | 13.8 (9.7--17.8) |
| ^y Includes only samples that have been mapped to individuals. @ Adjusted for sensitivities and specificities of RT-PCR, and antibody testing kits and procedures.^ | | | | |
